# Influence of *Schistosoma mansoni* infection on faecal calprotectin in the context of HIV, hepatitis B, and malaria co-infections

**DOI:** 10.1101/2025.09.16.25335894

**Authors:** Lauren Wilburn, Hanh Lan Bui, Paul Akampurira, Reagan Ddamba, David W. Oguttu, Betty Nabatte, Narcis B. Kabatereine, Goylette F. Chami

**Author notes:** Joint first authors (Lauren Wilburn, Hanh Lan Bui). These authors contributed equally to this work.

## Abstract

**Background:** The role of gut inflammation for intestinal schistosomiasis remains poorly understood in chronically infected and repeatedly treated populations.

**Methods:** We conducted a cross-sectional study nested in the SchistoTrack cohort within Pakwach district, Uganda. In 2024, 640 participants aged 6-85 years were examined for *Schistosoma mansoni* by Kato-Katz. fCal concentration was measured by ELISA. fCal was analysed as binary outcomes (detectable, ≥100 µg/g, >250 µg/g) and natural log-transformed continuous values. Co-endemic infections (malaria, HIV, hepatitis B (HBV)) and diverse sociodemographic covariates were investigated in logistic regressions with covariate selection. Correlations of symptoms, medications, and circulating white blood cells (WBC) with infections and fCal were reported.

**Results:** 74.4% of participants had detectable fCal, 22.3% had fCal ≥100 µg/g, and 7.0% had fCal >250 µg/g. *S. mansoni* prevalence was 49.1% (median 144 eggs per gram, IQR 36-369). Infection intensity was positively associated with all fCal outcomes (detectable: OR 1.20, ≥100 µg/g: OR 1.11, >250 µg/g: OR 1.26; continuous: β = 0.06) while status was positively related to all but the continuous fCal outcome. HIV was associated with fCal ≥100 µg/g (OR 2.52), while malaria and HBV were uninformative. Spearman analyses revealed significant and weak positive correlations between fCal, WBCs (neutrophils, monocytes and lymphocytes), faecal occult blood, medications (praziquantel and anti-inflammatories) and gastrointestinal symptoms (blood in stool).

**Conclusions:** *S. mansoni* infections are characterised by persistent, clinically concerning levels of gut inflammation in chronically infected populations with repeated praziquantel treatment. Integration of fCal thresholds into clinical guidelines may improve management of schistosomiasis-related morbidity.

## BACKGROUND

Intestinal schistosomiasis is a complex disease or set of conditions, where *Schistosoma mansoni* is the main causative species in sub-Saharan Africa (SSA) [1]. In endemic areas with high levels of schistosome re-exposure, chronic cases with a complex set of conditions of varying severity are observed. The most severe form is hepatosplenic schistosomiasis that includes periportal fibrosis (PPF) with hypersplenism, and ultimately complications of portal hypertension [2]. Less severe chronic presentations have been described with a focus on the non-specific outcomes of anaemia and diarrhoea [1]. The presence and persistence of chronic gut inflammation is less understood, and there remains a frequent misconception that repeatedly infected individuals are asymptomatic [3].

Parasite survival depends on successful egg translocation through the intestinal mucosa into the stool, where they are released into the environment to continue the life cycle within intermediate snail hosts found in freshwater [1]. Granulomas are necessary for this passage; however, uncontrolled immune responses and microulcerations caused by egg-induced tears in the intestinal epithelium may lead to localised gut inflammation [4]. Microulcerations caused by egg translocations may induce bleeding, and in severe cases, potential anaemia [5]. Yet, despite these less severe pathologies, the integrity of the gut lining is largely maintained, as there are few clinical cases of sepsis [5] – hypothesised to be due to the antimicrobial properties of schistosomes [6].

Faecal calprotectin (fCal) is a stable, non-specific biomarker of intestinal inflammation, widely used in the preliminary diagnosis and monitoring of clinically-relevant (i.e., requiring treatment) gut inflammation in inflammatory bowel diseases (IBD) such as Crohn’s disease and ulcerative colitis [7-9]. It is a calcium-binding protein released primarily by neutrophils and macrophages in response to local inflammation. Elevated fCal levels have been positively associated with a range of factors, including viral [10], parasitic [11], and bacterial [12] infections. In schistosomiasis-endemic settings, there is high co-endemicity of malaria, human immunodeficiency virus (HIV), and hepatitis B virus (HBV) [2]. Each has been independently linked to gut inflammation through different, and sometimes overlapping, hypothesised pathways, such as uncontrolled viral replication (HIV) [13], microvascular sequestration of parasitised red blood cells in the gut (malaria) [14], and changes to the gut microbiota (malaria, HIV, and HBV) [10, 14, 15]. Findings from the limited number of schistosomiasis-specific studies on fCal [11, 16, 17] primarily focus on young children, do not account for the influence of co-infections, and show inconsistent associations. Important questions remain regarding the persistence of gut inflammation in endemic populations with high re-infection, and the relevance in the context of repeated mass drug administration (MDA).

In this study, we explored the relationship between *S. mansoni* infection status and intensity with fCal levels in a nested, cross-sectional study within the community-based SchistoTrack cohort [18]. 640 individuals aged 6-85 were examined in 2024 in Pakwach district in Western Uganda. Co-endemic infections of malaria, HBV, and HIV were examined alongside current *S. mansoni* infection status and intensity. Our aim was to identify whether gut inflammation occurs within the context of chronically and repeatedly infected *S. mansoni* populations with a repeated history of praziquantel treatment through MDA, and whether this relationship is attenuated by co-infections.

## METHODS

### Study context and participant sampling

A cross-sectional study was conducted in Pakwach district within the SchistoTrack cohort, where >50% *S. mansoni* prevalence has been observed [2, 18, 19]. SchistoTrack examines liver disease progression and related multimorbidity with annual follow-ups since 2022. As of 2024, Pakwach had at least fifteen annual rounds of school-based and community-based MDA. At recruitment for SchistoTrack, one child aged five years or older and one adult aged 18 years or older were selected within each randomly sampled household. Detailed SchistoTrack sampling is described elsewhere [18]. In 2024, 750 individuals from the first 12 study villages visited in Pakwach were targeted to oversample the population needed for study, as several exclusion criteria were applied that focused on the completeness of exposures (Figure 1).

**Figure 1.**
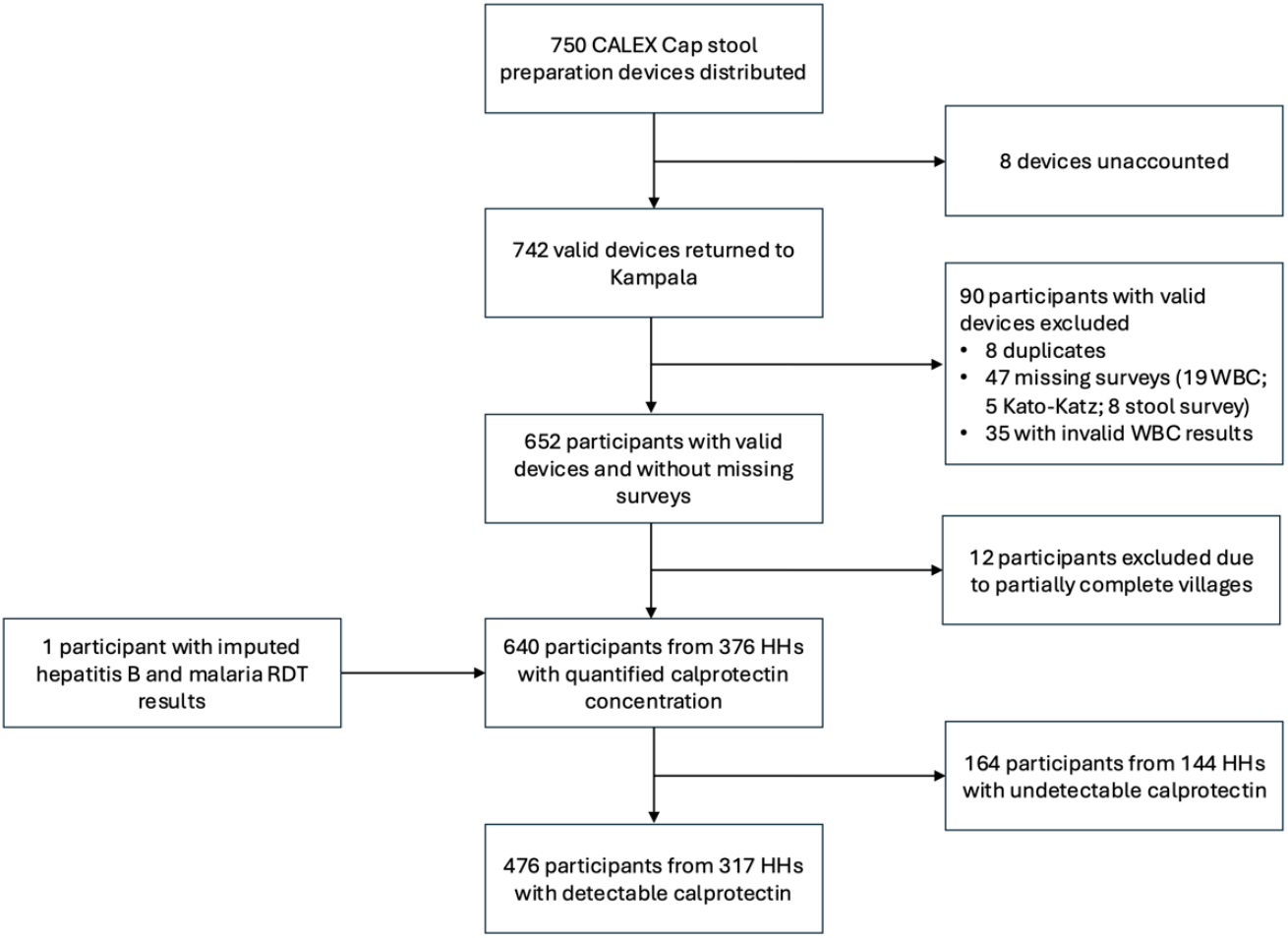
Participant flow diagram.

### Outcomes

fCal concentration was measured using commercial ELISA kits (Bühlmann Laboratories AG) as a continuous concentration in µg/g of stool (Supplementary Methods). Three fCal outcomes were constructed for analysis. First, a binary outcome was defined as detectable fCal and coded as one if the concentration was greater than or equal to the detection limit, otherwise zero. In clinical practice in high-income settings, a traffic light system based on fCal concentration is commonly used as a triage tool for suspected IBD or clinically relevant gut inflammation [8, 9]. Calprotectin levels lower than 100 µg/g indicate low IBD risk, values between 100-250 µg/g indicate an intermediate risk, while values above 250 µg/g indicate high risk [8, 9]. To examine the clinical relevance of these thresholds for *S. mansoni* infection, two additional binary outcomes were created based on ≥100 µg/g and >250 thresholds. To explore the relevance of fCal for schistosomiasis without relying on pre-defined clinical cut-offs, fCal was treated as a continuous variable and natural log-transformed to approximate a normal distribution. Individuals with values below the detection limit were excluded for this outcome, as their exact values were unknown but possibly non-zero.

### Exposures

The primary exposure was *S. mansoni* infection intensity and status, as determined by Kato-Katz microscopy [20]. Infection intensity was analysed as the natural log-transformed eggs per gram (EPG), and positive infection status was defined as EPG ≥1. Secondary exposures included malaria parasite density and *Plasmodium falciparum* infection status. Malaria parasite density (parasites/µL of blood, not species-specific) was measured using microscopy of thick smear slides, while infection status was determined by rapid diagnostic testing (RDT) (Abbott Bioline Malaria Ag P.f/Pan) [21]. HIV status was determined by a series of RDTs following the Uganda national testing algorithm (Abbott Determine HIV-1/2, Chembio Diagnostics HIV-1/2 STAT-PAK Assay, Abbott Bioline HIV 1/2 3.0) [22]. HBV status was determined by surface antigens detected using the Abbott Determine HBsAg 2 test. HBV infection chronicity was defined as being HBV-positive in 2023 and 2024 among those who attended both follow-up timepoints in SchistoTrack. Infection status and HBV chronicity were coded as binary variables, while *S. mansoni* infection intensity and malaria parasite density were treated as continuous variables. To explore clinical, mechanistic and confounding relationships with exposures and fCal, white blood cell (WBC) differentials, faecal occult blood (FOB), and haemoglobin (Hb) counts were measured, as well as gastrointestinal symptoms and medications relevant to gut inflammation and infections. Viral loads were measured for people living with HIV (PLHIV). Detailed descriptions of biomedical variables are provided in the Supplementary Methods.

### Sociodemographic covariates

Individual-level covariates included age, gender, and occupation (farmer, fisherman, fishmonger, and none/other as a reference level). Household-level covariates included household size, home quality score, years of residence in the village, home ownership status, social status, year of recruitment, main type of healthcare facility used by the household, and household distance to the nearest government health centre. Other household-level covariates included improved sanitation facility at home, improved drinking water source, and household treated drinking water. Detailed definitions and collection procedures are provided elsewhere [18].

### Statistical analysis

Analyses were run in R (v4.2.1) [23]. Logistic regression models for binary fCal outcomes and linear regression model for the continuous natural log-transformed fCal outcome were specified through backwards stepwise model selection using the lowest Bayesian information criterion (BIC) to produce parsimonious interpretable models [23]. *S. mansoni* infection intensity, malaria parasite density, HIV status and HBV status were core variables forced into all models. We also examined *S. mansoni* and malaria infection status in place of intensity and density measures. To investigate potential mediators of fCal, natural log-transformed neutrophil count (continuous) and FOB (binary) were added to the models. Detailed analyses are described in the Supplementary Methods.

## RESULTS

### fCal variation

640 of participants aged 6-85 from 376 households with complete data had fCal measured (Figure 1). Those participants were representative of the broader SchistoTrack cohort (Table S1). 74.38% (476/640) of participants had detectable levels of fCal. The median detectable concentration was 54.77 µg/g (IQR 25.69-123.66). fCal had a left-skewed distribution (skewness 6.18, kurtosis 44.12) (Figure 2A). 22.34% (143/640) of participants (including those with undetectable fCal) had fCal concentration ≥100 µg/g, while 7.03% (45/640) had fCal concentration >250 µg/g. Across age, fCal was slightly elevated in young children (6-10 years) and in older adults (>60 years), although only 6.56% (42/640) of participants were over 60 (Figure 2B).

**Figure 2.**
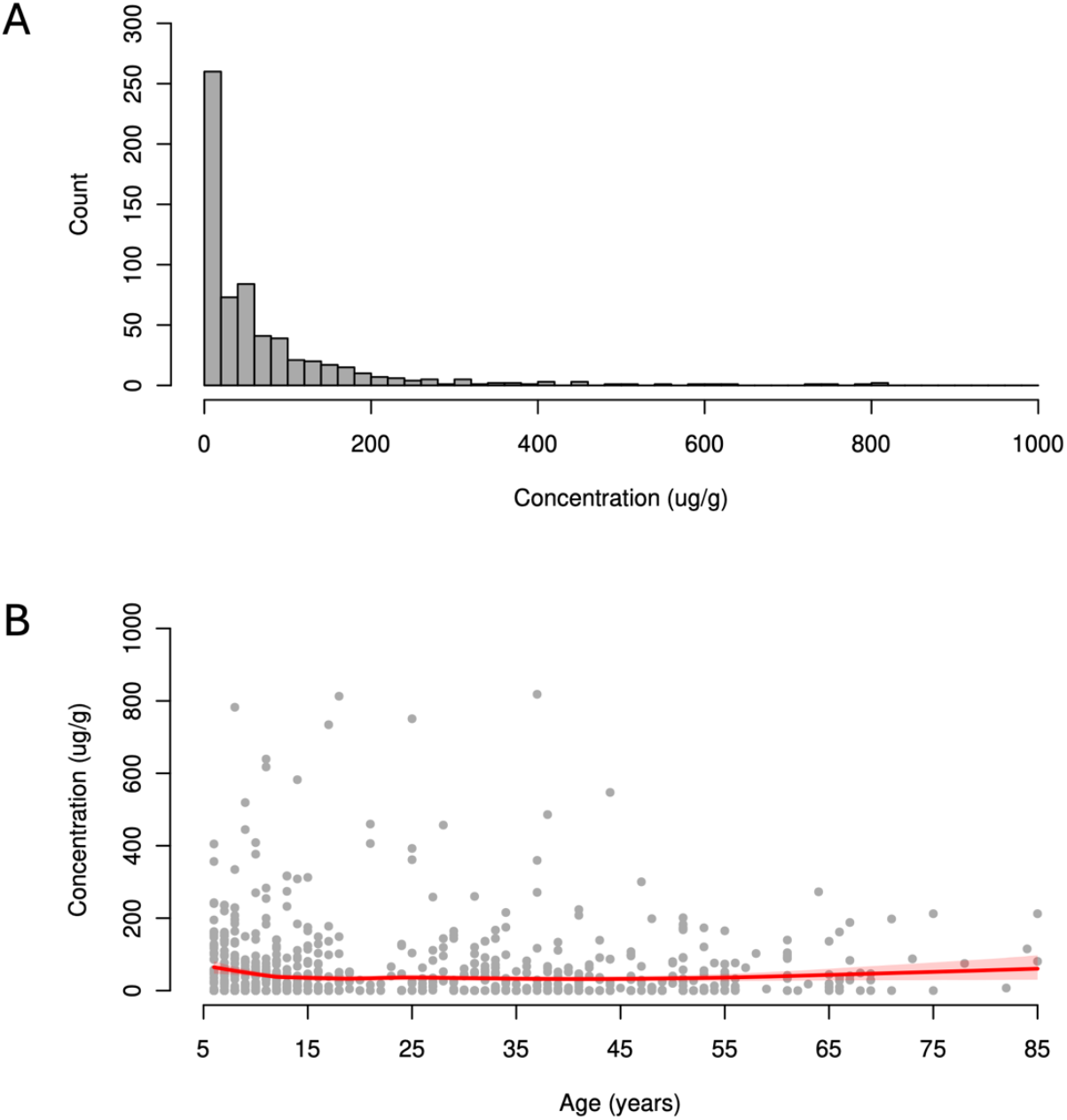
Calprotectin concentration among study participants (n = 631). (A) Histogram of calprotectin concentration. (B) Scatter plot of calprotectin concentration over age with a LOWESS curve (smoothing span = 0.5; red line with shaded areas representing bootstrap 95% confidence intervals). 9 participants with calprotectin concentrations >1000 µg/g (range: 1490-2290 µg/g) were excluded from the plots.

### Infection prevalence

Participant characteristics are presented in Table 1. 49.06% (314/640) of the study participants were infected with *S. mansoni*, with 23.89% (75/314) of those infected classified as having high infection intensity (≥400 EPG). Among those *S. mansoni*-positive, the median EPG was 144 (IQR 36-369). 36.72% (235/640) of participants had malaria. Among those with available microscopy results (67.66%, 159/235), 94.34% (150/159) of infections were caused by *P. falciparum*. Among those with malaria, the median parasite density was 48 parasites/µL of blood (IQR 0-336). 5.31% (34/640) of participants were HBV positive. Among the HBV positive participants, 85.29% (29/34) had been tested for HBV in 2023, of whom 75.86% (22/29) were chronically infected (positive in both 2023 and 2024). 4.84% (31/640) of participants were living with HIV, 83.87% (26/31) of whom were aware of their status, and all of those (100%, 26/26) were currently on antiretroviral therapy (ART). Among participants infected with *S. mansoni*, the co-infection prevalence was 5.10% (16/314) for HBV, 3.18% (10/314) for HIV, and 44.90% (141/314) for malaria. One participant was co-infected with HBV and HIV.

**Table 1.** Participant characteristics.

|  | Overall (N=640) | Detectable fCal (N=476) | Undetectable fCal (N=164) |
| --- | --- | --- | --- |
| *S. mansoni* status (positive) | 314 (49.1%) | 256 (53.8%) | 58 (35.4%) |
| *S. mansoni* intensity (EPG) | 0 (0–144) | 12 (0–192) | 0 (0–24) |
| Malaria status (positive) | 235 (36.7%) | 178 (37.4%) | 57 (34.8%) |
| Malaria parasite density | 0 (0–0) | 0 (0–3.9) | 0 (0–0) |
| HIV status (positive) | 31 (4.8%) | 25 (5.3%) | 6 (3.7%) |
| HBV status (positive) | 34 (5.3%) | 26 (5.5%) | 8 (4.9%) |
| Age | 19 (11–40) | 18 (10.75–39) | 26 (11.75–43) |
| Gender (female) | 328 (51.3%) | 245 (51.5%) | 83 (50.6%) |
| Occupation |  |  |  |
| Farmer | 103 (16.1%) | 79 (16.6%) | 24 (14.6%) |
| Fisherman | 30 (4.7%) | 23 (4.8%) | 7 (4.3%) |
| Fishmonger | 6 (0.9%) | 5 (1.1%) | 1 (0.6%) |
| None/other | 501 (78.3%) | 369 (77.5%) | 132 (80.5%) |
| Education |  |  |  |
| None | 124 (19.4%) | 94 (19.7%) | 30 (18.3%) |
| Primary | 471 (73.6%) | 347 (72.9%) | 124 (75.6%) |
| Secondary or above | 45 (7.0%) | 35 (7.4%) | 10 (6.1%) |
| Year of recruitment |  |  |  |
| 2024 | 64 (10.0%) | 46 (9.7%) | 18 (11.0%) |
| 2023 | 470 (73.4%) | 329 (69.1%) | 141 (86.0%) |
| 2022 | 106 (16.6%) | 101 (21.2%) | 5 (3.0%) |
| No. of people in household | 3 (3–4) | 3 (3–4) | 3 (3–4) |
| Home quality | 3 (3–3) | 3 (3–3) | 3 (3–3) |
| Social status | 108 (16.9%) | 80 (16.8%) | 28 (17.1%) |
| Home ownership | 586 (91.6%) | 431 (90.5%) | 155 (94.5%) |
| Years lived in village | 17.5 (8.0–34.3) | 17.5 (8.0–35) | 17.5 (7.8–33.3) |
| Distance to the nearest gov't health centre | 6.4 (5.2–12.2) | 6.4 (3.9–12.2) | 6.6 (5.8–12.1) |
| Main type of health facility used |  |  |  |
| Private clinic or other | 43 (6.7%) | 37 (7.8%) | 6 (3.7%) |
| Gov't health centre | 597 (93.3%) | 439 (92.2%) | 158 (96.3%) |
| Improved drinking water source | 331 (51.7%) | 218 (45.8%) | 113 (68.9%) |
| Improved sanitation facility at home | 307 (48.0%) | 249 (52.3%) | 58 (35.4%) |
| Household treated drinking water | 173 (27.0%) | 136 (28.6%) | 37 (22.6%) |
| Any intestinal symptoms | 251 (39.2%) | 184 (38.7%) | 67 (40.9%) |
| Any abdominal pain | 199 (31.1%) | 138 (29.0%) | 61 (37.2%) |
| Any abdominal enlargement | 20 (3.1%) | 17 (3.6%) | 3 (1.8%) |
| Vomiting or nausea | 17 (2.7%) | 15 (3.2%) | 2 (1.2%) |
| Faecal occult blood | 128 (20.0%) | 101 (21.2%) | 27 (16.5%) |
| Any blood in stool | 50 (7.8%) | 44 (9.2%) | 6 (3.7%) |
| Blood in stool with diarrhoea | 23 (3.6%) | 21 (4.4%) | 2 (1.2%) |
| Blood in stool without diarrhoea | 33 (5.2%) | 30 (6.3%) | 3 (1.8%) |
| Melena | 5 (0.8%) | 4 (0.8%) | 1 (0.6%) |
| Any diarrhoea without blood | 185 (28.9%) | 143 (30.0%) | 42 (25.6%) |
| Diarrhoea (reported) | 37 (5.8%) | 33 (6.9%) | 4 (2.4%) |
| Diarrhoea (Bristol stool scale) | 159 (24.8%) | 120 (25.2%) | 39 (23.8%) |
| Constipation (Bristol stool scale) | 64 (10.0%) | 48 (10.1%) | 16 (9.8%) |
| Anaemia | 371 (58.0%) | 267 (56.1%) | 104 (63.4%) |
| Haemoglobin count (g/dL) (1 missing) | 11.4 (9.8–13.2) | 11.6 (9.8–13.2) | 11.3 (9.8–13.0) |
| Praziquantel *Past year* | 519 (81.1%) | 386 (81.1%) | 133 (81.1%) |
| MDA | 25 (3.9%) | 16 (3.4%) | 9 (5.5%) |
| Study | 514 (80.3%) | 384 (80.7%) | 130 (79.3%) |
| Other deworming medications *Past month* | 55 (8.6%) | 40 (8.4%) | 15 (9.1%) |
| Antimalarials *Past month* | 202 (31.6%) | 143 (30.0%) | 59 (36.0%) |
| Antibiotics *Past month* | 294 (45.9%) | 217 (45.6%) | 77 (47.0%) |
| Anti-inflammatories *Past month* | 37 (5.8%) | 22 (4.6%) | 15 (9.1%) |
| HIV ART *Ever* | 26 (4.1%) | 22 (4.6%) | 4 (2.4%) |
| HBV ART *Ever* | 0 (0.0%) | 0 (0.0%) | 0 (0.0%) |
| Total WBC count | 5.9 (4.5–7.53) | 6.15 (4.7–7.73) | 5.3 (4.1–6.7) |
| Neutrophil count | 2.1 (1.5–2.9) | 2.15 (1.6–3) | 1.9 (1.4–2.6) |
| Monocyte count | 0.4 (0.3–0.5) | 0.4 (0.3–0.5) | 0.3 (0.2–0.4) |
| Lymphocyte count | 2.9 (2.1–3.7) | 3 (2.2–3.9) | 2.7 (2–3.4) |
| Eosinophil count | 0.3 (0.1–0.6) | 0.3 (0.1–0.6) | 0.3 (0.1–0.5) |
| Basophil count | 0 (0–0) | 0 (0–0) | 0 (0–0) |
| Neutrophil-to-lymphocyte ratio (2 missing) | 0.73 (0.55–0.96) | 0.74 (0.55–0.96) | 0.70 (0.54–0.97) |

Data are presented as N (%) or median (IQR).

### Determinants of fCal

Comparison of participants with detectable and undetectable fCal are shown in Table 1. Unadjusted analyses are shown in Tables S2-S5. *S. mansoni* infection intensity was positively associated with all fCal outcomes in adjusted models (Figure 3). Each 2.72-fold increase in EPG was associated with 1.20 times higher odds of having detectable fCal (95% CI 1.10-1.30) and 1.11 times higher odds of fCal ≥100 µg/g (95% CI 1.04-1.20). The association was stronger for a higher fCal threshold of >250 µg/g (OR 1.26, 95% CI 1.12-1.42). In a linear model, a 10% increase in EPG was associated with a 0.57% increase in fCal (95% CI 0.19-0.96%). HIV status was associated with 2.52 (95% CI 1.18-5.40) times higher odds of fCal ≥100 µg/g, although unrelated to high levels of gut inflammation (fCal >250 µg/g) (Figure 3B, 3C). Co-infection with HIV and *S. mansoni* was positively correlated with fCal ≥100 µg/g (Figure S1). Malaria and HBV were not significantly associated with any fCal outcomes. Among participants who attended both 2023 and 2024 follow-up timepoints, chronic HBV infection was also not correlated with any fCal outcomes. Female participants had 3.05 (95% CI 1.52-6.10) times higher odds of fCal >250 µg/g than male participants; age was not selected in any fCal model (Figure 3). Access to an improved drinking water source was associated with 0.48 (95% CI 0.24-0.93) times lower odds of having detectable fCal compared to unimproved source (Figure 3A). Improved sanitation facility and year of recruitment were selected in the model but not significant (Figure 3A). When *S. mansoni* infection intensity and malaria parasite density were replaced with infection status, *S. mansoni* associations remained significant for detectable fCal and low (≥100 µg/g) and high (>250 µg/g) inflammation levels, but were borderline insignificant for linear fCal (Figure S2). Results remained robust when one participant with imputed malaria and HBV status was excluded from the models.

**Figure 3.**
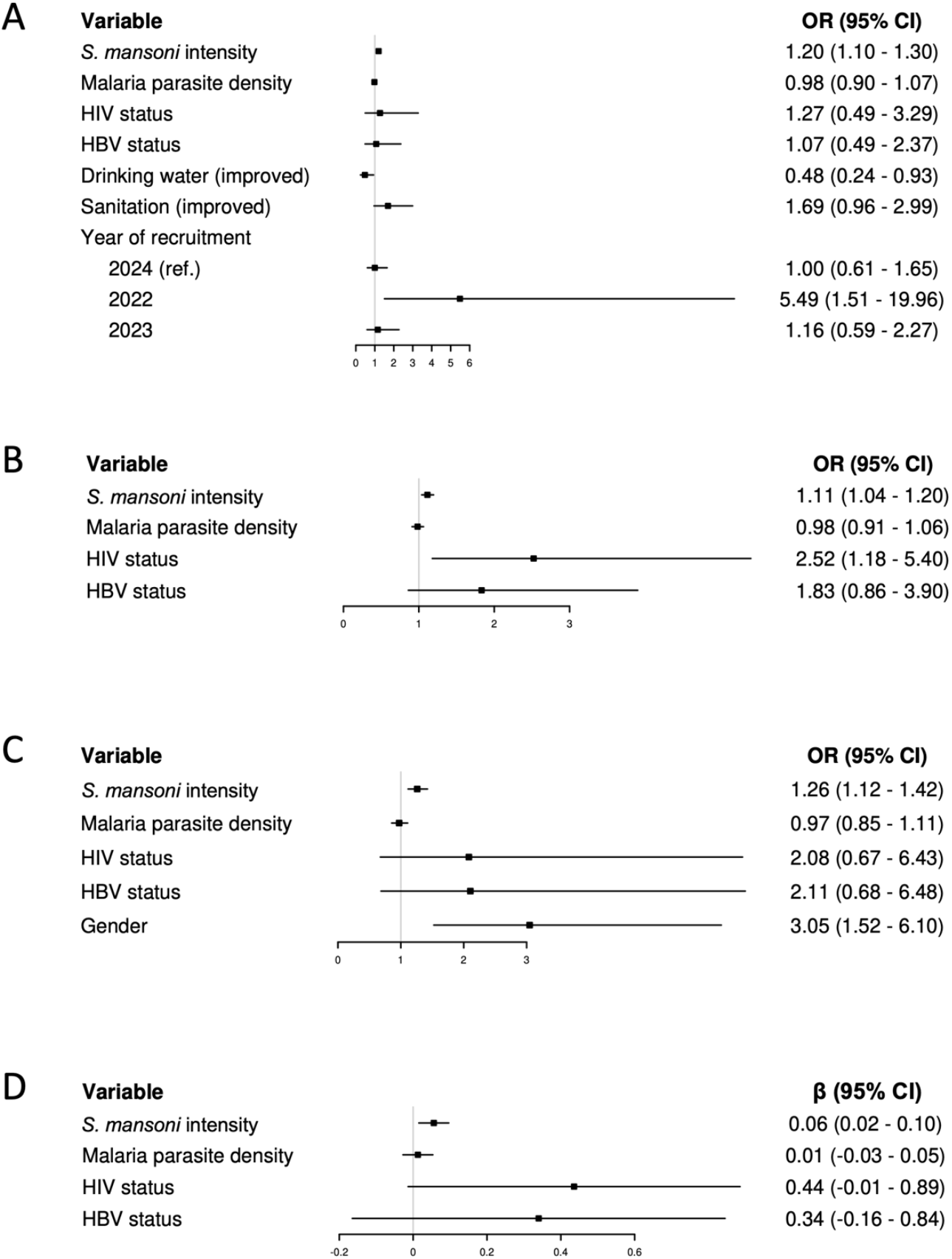
Calprotectin (fCal) models for infection intensity. Model of (A) detectable fCal, (B) ≥100 µg/g, (C) fCal >250 µg/g, and (D) linear fCal (N = 640). Logistic regression models were selected by backward stepwise selection based on the lowest Bayesian Information Criterion. 95% confidence intervals were calculated using clustered standard errors at the village level (for detectable fCal; number of villages = 12) and the household level (for linear fCal; number of households = 376). Floating absolute risks were calculated for the year of recruitment variable. ICC (village) = 0.147; 0.029; 0.014; 0.016. ICC (household) = 0.119; 0.062; 0.084; 0.102. VIF range: 1.00-1.09; 1.00-1.04; 1.01-1.14; 1.00-1.04. AUC for stratified ten-fold cross-validation = 0.69; 0.57; 0.70. R^2^ (linear model) = 0.03.

### WBC differentials and fCal production

Total WBC, neutrophil and lymphocyte count were weakly positively correlated with detectable fCal (r_s_ 0.14, *p* < 0.05; r_s_ 0.13, *p* < 0.05; r_s_ 0.12, *p* < 0.05) (Figure 4A). Monocyte count was weakly positively correlated with detectable fCal and both fCal thresholds (r_s_ 0.12, *p* < 0.05 for detectable fCal; r_s_ 0.09, *p* < 0.05 for fCal ≥100 µg/g; r_s_ 0.09, *p* < 0.05 for fCal >250 µg/g), while eosinophil percentage was positively correlated only with fCal >250 µg/g (r_s_ 0.08, *p* < 0.05). All WBC differentials except for basophils were weakly to moderately positively correlated with *S. mansoni* infection intensity and status (r_s_ range 0.13-0.22, *p* < 0.05). The neutrophil-to-lymphocyte ratio was not significantly correlated with fCal or *S. mansoni* outcomes (r_s_ range −0.06-0.06, *p* ≥ 0.05). When added to the adjusted models, the natural logarithm of the neutrophil count was associated with 1.96 (95% CI 1.13-3.40) times higher odds of having detectable fCal only, where *S. mansoni* infection intensity and status remained significant (Figures S3, S4).

**Figure 4.**
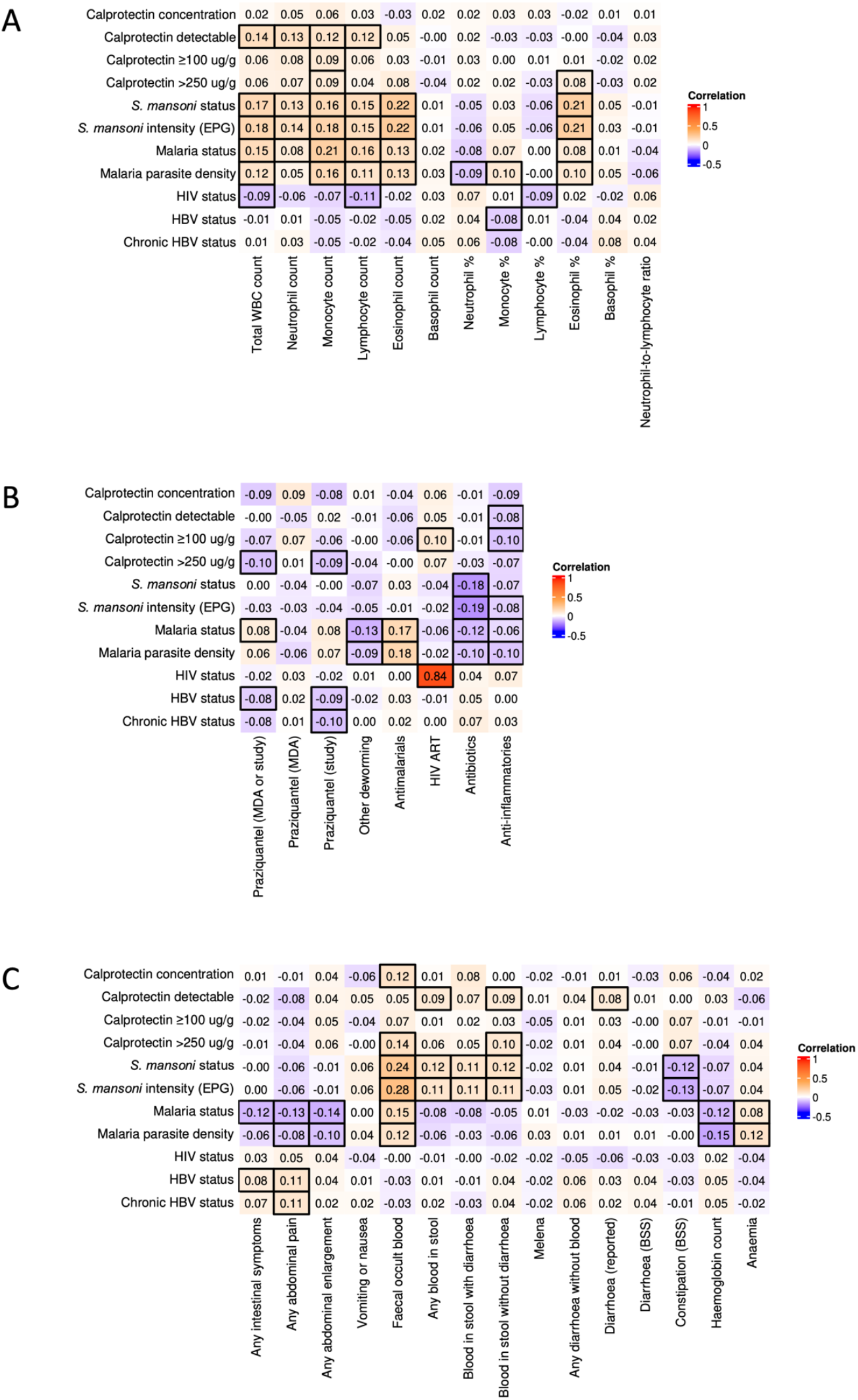
Spearman correlation matrices of (A) white blood cell differentials, (B) medications, and (C) symptoms with fCal outcomes and infections (*S. mansoni*, malaria, HIV and HBV).

### Medications as potential fCal confounders

Praziquantel taken within the past year (through MDA or the study) was negatively correlated only with high levels of gut inflammation (fCal >250 µg/g) (r_s_ −0.10, *p* < 0.05) (Figure 4B). Praziquantel slightly attenuated the effect of *S. mansoni* status in the adjusted model (OR 1.99, 95% CI 1.01-3.95), while it was not significant in the fCal models with *S. mansoni* intensity (Figure S2C, 3C). Ever taking ART for HIV was positively correlated with fCal ≥100 µg/g (r_s_ 0.10, *p* < 0.05) (Figure 4B), but non-significant when added to the adjusted model (Figure 3B). 83.87% (26/31) of PLHIV had viral load measured in 2025, of which only 50% (13/26) were virally suppressed. Among PLHIV only, viral suppression was not significantly correlated with fCal. Taking anti-inflammatory medications was negatively correlated only with detectable fCal (r_s_ −0.08, *p* < 0.05) and fCal ≥100 µg/g (r_s_ −0.10, *p* < 0.05). Anti-inflammatory medications also were negatively correlated with *S. mansoni* infection intensity (r_s_ −0.08, *p* < 0.05), while antibiotics were negatively correlated with *S. mansoni* infection intensity (r_s_ −0.19, *p* < 0.05) and status (r_s_ −0.18, *p* < 0.05) but not fCal outcomes. In the adjusted models, anti-inflammatory medications were associated with 0.18 (95% CI 0.04-0.77) times lower odds of fCal ≥100 µg/g, and a 31.61% decrease in fCal levels in the linear model (95% CI −51.32% to −2.96%) (Figure 3B, 3D). *S. mansoni* infection intensity remained significant in both models. *S. mansoni* status was no longer significant in the fCal ≥100 µg/g model, while it remained non-significant in the linear model. Anti-inflammatory medications were associated with 0.56 (95% CI 0.34-0.93) times lower odds of having detectable fCal when added to infection status models only (Figure 3A), where *S. mansoni* status remained significant.

### Microulcerations detected by FOB and gastrointestinal symptoms

FOB was weakly positively correlated with fCal concentration (r_s_ 0.12, *p* < 0.05) and fCal >250 µg/g (r_s_ 0.14, *p* < 0.05) (Figure 4C). FOB also was moderately positively correlated with both *S. mansoni* infection intensity (r_s_ 0.28, *p* < 0.05) and status (r_s_ 0.24, *p* < 0.05). When added to the adjusted models, FOB was associated with 2.96 (95% CI 1.48-5.94) times higher odds of fCal >250 µg/, and a 0.30% increase in fCal concentration in the linear model (95% CI 0.08-0.53%) (Figure S5). *S. mansoni* infection intensity remained significant in the fCal >250 µg/g model, while it became borderline non-significant in the linear model (Figure S5). *S. mansoni* status was insignificant in both FOB models (Figure S6).

Having any blood in stool (with or without diarrhoea) within the past month was weakly positively correlated with detectable fCal only (r_s_ 0.09, *p* < 0.05), while having blood in stool without diarrhoea was weakly positively correlated with detectable fCal (r_s_ 0.09, *p* < 0.05) and fCal >250 µg/g (r_s_ 0.10, *p* < 0.05) (Figure 4C). Both symptoms were weakly positively correlated with *S. mansoni* infection intensity (r_s_ 0.11, *p* < 0.05; r_s_ 0.11, *p* < 0.05) and status (r_s_ 0.12, *p* < 0.05; r_s_ 0.12, *p* < 0.05). Having blood in stool with diarrhoea within the past month was weakly positively correlated with *S. mansoni* infection intensity (r_s_ 0.11, *p* < 0.05) and status (r_s_ 0.11, *p* < 0.05) but not fCal outcomes. Reported diarrhoea within the past month was positively correlated with detectable fCal only (r_s_ 0.08, *p* < 0.05), while constipation based on biosample analysis was negatively correlated with *S. mansoni* intensity (r_s_ −0.13, *p* < 0.05) and status (r_s_ −0.12, *p* < 0.05) but not fCal outcomes. Hb count and anaemia were not correlated with fCal outcomes or *S. mansoni* infection intensity and status (r_s_ range −0.07-0.04, *p* ≥ 0.05). Only one participant reported vomiting blood.

## DISCUSSION

No World Health Organization guidelines based on clinical biomarkers exist for schistosomiasis morbidity. Here we investigated the relevance of fCal – a clinically-established, non-invasive marker of gut inflammation – for infection with *S. mansoni*, while considering important co-endemic infections of HIV, HBV, and malaria, alongside WBC differentials, medications, and symptoms. We studied 640 individuals aged 6-85 years within the community-based SchistoTrack cohort, and found consistent associations between *S. mansoni* infection status and intensity with elevated fCal levels as well as evidence of microulcerations contributing to this association.

*S. mansoni* infection – both intensity and status – were positively associated with fCal outcomes of varying severity, despite our study area having received fifteen rounds of annual MDA. Infection intensity was significant for all fCal outcomes, while infection status was significant for all outcomes except for linear fCal, where it was borderline non-significant. These findings support the potential relevance of fCal as an inflammatory biomarker for *S. mansoni*, even when applying established clinical thresholds for IBD, where fCal ≥100 µg/g indicates intermediate risk and fCal >250 µg/g indicates high risk of IBD [8]. The association between *S. mansoni* status and gut inflammation, given chronic exposure in our study population, suggests that a high level of morbidity may persist in current infection. Surprisingly, age was not relevant in most fCal models, despite prior studies that focused exclusively on children and did not account for co-infections [11, 16, 17]. This finding raises the possibility that gut inflammation continues even in the presence of acquired immunity from repeated schistosome exposure. More detailed immunological investigations of the gut, such as proteomic analyses, are needed, alongside further epidemiological studies to understand the relationship between gut inflammation and sepsis beyond murine models [5].

Praziquantel treatment was negatively associated with high levels of gut inflammation (fCal >250 µg/g) and very minimally attenuated the effect of *S. mansoni* status on fCal. Notably, praziquantel had no significant effect on the relationship between infection intensity and gut inflammation. This suggests that although praziquantel effectively reduces infection intensity, it may not resolve established intestinal inflammation – likely because it has limited or varied effect on intact granulomas. Evidence from a murine model [24] supports this interpretation, showing that the impact of praziquantel on hepatic and intestinal granuloma volume depends on both dose and treatment duration, with shorter or single high-dose regimens being associated with smaller or no reductions.

FOB showed a positive correlation with both fCal and *S. mansoni* and slightly attenuated the effect of *S. mansoni* intensity. Microulcerations and associated granulomatous inflammation in the gut epithelium may be caused by *S. mansoni* egg translocation and entrapment, and could be a major factor in clinically-relevant gut inflammation [25]. Neither fCal or *S. mansoni* were correlated with anaemia or haemoglobin count, suggesting a lack of severe complications due to intestinal bleeding, and questioning the use of conventional symptoms for schistosomiasis morbidity representation. Our findings extend beyond previous studies that have only linked FOB to schistosome infection [16, 17].

Higher neutrophil counts were linked to detectable fCal and *S. mansoni*, while monocytes were associated with all fCal outcomes and *S. mansoni*. Neutrophils, which contain high levels of calprotectin, act as early responders in acute inflammation, releasing calprotectin to recruit other immune cells, including monocytes [26]. Their contribution to chronic inflammation is emerging and may involve sustained immune activation and tissue damage [27]. The exact role of neutrophils in host immune response to *S. mansoni* infection is not well understood [28], as research has predominantly focused on eosinophils (a positive association also observed in our study). Our findings suggest circulating neutrophils contribute to gut inflammation in *S. mansoni* infection and highlight WBC differentials as useful indicators of immune response. Further research is needed to establish the clinical relevance of neutrophilia in schistosomiasis and compare circulating with gut-localised WBCs. Neutrophils also may reflect fCal release in response to acute inflammation from other infections.

In our study, HIV status was positively associated with fCal ≥100 µg/g. Although effective ART achieves viral suppression and improves life expectancy of PLHIV, morbidity remains due to chronic immune activation and inflammation [29]. The gut is a primary site of HIV replication, and gut inflammation is a hallmark of infection [29]. During acute infection, CD4^+^ T-lymphocyte depletion and loss of gut integrity trigger gut inflammation, microbial translocation and dysbiosis, and subsequent systemic inflammation and immune activation, which persist at residual levels even with successful ART [10, 30]. The association of both *S. mansoni* and HIV with fCal, and the positive correlation between co-infection and fCal, underscores the need to investigate the detailed contribution of co-infections to gut inflammation, given the role of HIV and schistosome infection already shown in the liver [2]. Recent research has shown that elevated fCal in ART-treated PLHIV is linked to systemic inflammation, suggesting an important area for future research on HIV-schistosome co-infections [10].

Self-reported symptoms such as blood in stool and diarrhoea are frequently used for assessing acute *S. mansoni* infection [1]. In our study, blood in stool was weakly positively correlated with both fCal and *S. mansoni* infection, possibly indicating intestinal blood loss due to microulcerations [25]. *S. mansoni*, despite representing chronic infection or repeated exposure within our study population, remained (weakly) associated with a wide range of gastrointestinal symptoms, suggesting that symptoms should be used not only in acute but also chronic infection monitoring for schistosomiasis. In frequently re-exposed populations, chronic morbidity and the associated disability-adjusted life years may be underestimated if clinical manifestations are not fully captured in health states or disability weights [31-33]. Furthermore, our finding that gut inflammation persists in repeatedly treated populations raises questions about how ongoing subclinical morbidity contributes to severe *S. mansoni*-associated morbidities, including end-stage periportal fibrosis [2].

The strengths of this study lie in the comprehensiveness of co-infections, symptoms, and potential confounders observed across a wide age range of individuals. Concerning limitations, we did not collect data on concurrent bacterial infections, which may also influence fCal levels [12]. However, we attempted to partially account for bacterial effects by incorporating higher-level environmental factors, such as improved sanitation facility, improved drinking water source, and household treated drinking water. These factors were generally not selected in our models, although individual-level hygiene behaviour data were not available. Laboratory analyses were performed using a single sensitivity threshold without re-analysis at lower thresholds, and each sample was tested in a single replicate, potentially limiting measurement precision. Data were collected at a single time point, providing only a cross-sectional snapshot.

We showed that intestinal schistosomiasis is characterised by gut inflammation, particularly in chronically infected populations where significant morbidity due to *S. mansoni* persists despite repeated MDA. The observed link between *S. mansoni* infection and elevated fCal supports its potential role as a practical, non-invasive biomarker for gut inflammation in intestinal schistosomiasis. Incorporating such measures to develop schistosomiasis morbidity or clinical management guidelines could improve patient management within primary healthcare facilities beyond controlling infection alone.

## Data Availability

Individual participant data is not available due to the identifiable nature of the participant characteristics and ongoing nature of the cohort. Extensive metadata relevant to the study are included in the article or uploaded as supplementary information. No new model code was developed for the analyses in this study.

## DECLARATIONS

### Author contributions

GFC accepts full responsibility for the finished work and/or the conduct of the study, had access to the data, and controlled the decision to publish. Conceptualisation: GFC. Data curation: LW, PA, RD, DWO, BN, and GFC. Formal analysis: LW and HLB. Funding acquisition: GFC. Investigation: LW, HLB, and GFC. Methodology: LW, HLB, and GFC. Project administration: NBK and GFC. Resources: NBK and GFC. Software: GFC. Supervision: GFC. Validation: HLB. Visualisation: HLB. Writing the original draft: HLB and GFC. Writing-review and editing: LW, HLB, PA, RD, DWO, BN, NBK, and GFC.

### External meeting

This work has not been presented at an external meeting.

### Ethics approval

This study involves human participants. Data collection and use were reviewed and approved by the Oxford Tropical Research Ethics Committee (OxTREC 509-21), the Vector Control Division Research Ethics Committee of the Uganda Ministry of Health (VCDREC146) and the Uganda National Council of Science and Technology (UNCST HS 1664ES). Written informed consent was obtained for adult participants, with adults consenting on behalf of the children after receiving their informed assent. Ethics approval was obtained from all local ethics committees. GFC is the principal investigator of SchistoTrack. The principal investigator’s institution is the University of Oxford, Oxford, UK. Participants gave informed consent to participate in the study before taking part.

## Acknowledgements

We thank the SchistoTrack Group, especially Christin Puthur, for their valuable feedback. We also give most thanks to the field teams, including surveyors, technicians, nurses, sonographers, malacologists and auxiliary workers who are part of SchistoTrack. A special acknowledgement is due to the study participants, village health team members, local government nurses and surveyors and district leadership.

