## Supplementary material for "Influence of *Schistosoma mansoni* infection on faecal calprotectin in the context of HIV, hepatitis B, and malaria co-infections"

### Supplementary Methods

#### Calprotectin (fCal) quantification

The first stool of the day was collected. CALEX Cap (Bühlmann Laboratories AG, Schönenbuch, Switzerland) were used for sample storage and protein extraction. Samples were vortexed and stored in a cool box in the field and during transportation. Samples were transported within one week from initial preparation to the Vector Control Division, Kampala, where they were stored at -20°C until use. Faecal fCal concentration was quantified using fCal ELISA kits (Bühlmann Laboratories AG, Schönenbuch, Switzerland) according to manufacturer instructions. In brief, frozen samples were defrosted at room temperature for at least 30 minutes prior to use and diluted 1:150 in an assay buffer. In duplicate, 100 µL of blanks, calibrators, and controls were eluted into a 96-well fCal ELISA plate. Single replicates of the samples were run. Plates were covered and incubated at room temperature, with agitation at 450 rpm for 35 minutes. Wells were washed three times with wash buffer, and 100 µL of enzyme label was added to all wells and incubated for an additional 30 minutes. Wells were washed five times with a wash buffer, and 100 µL of substrate solution was added and incubated for 15 minutes. Subsequently, 100 µL of stop solution was added, and the absorbance was read at 450 nm on a microtiter plate reader (Agilent, Biotek) and with associated microplate reader and imager software, Gen 5 (Aligent, BioTek). Data was exported to Microsoft Excel, and calibration curves were constructed in R. The final concentration of fCal was expressed as µg/g of stool. The limit of detection was 12.6 µg/g and the limit of quantification was 30 µg/g. Due to the interest in increased inflammation levels and limited supply of assays, samples that fell below the detectable limit were not re-run at more sensitive levels.

#### Statistical analysis

Individual and household-level covariates were allowed to be selected in BIC-based models, as there were no prior assumptions on biological importance. Variable selection was performed for each fCal outcome separately. Standard errors were clustered if the intraclass correlation coefficient (ICC) was ≥0.10 for either village or household (using the higher of the two) [1, 2]. ICC was calculated for assay plate but not used for clustering, as samples from the same village were usually run on a single plate. Floating absolute risks were calculated for the selected categorical variables to assess the chosen Ref level [3]. Collinearity was assessed using variance inflation factors (VIFs) [4]. The mean area under the receiver operating characteristic (ROC) curve (AUC) across stratified ten-fold cross-validation was calculated as a measure of predictive performance [5]. Spearman correlation matrices were used to explore relationships between white blood cell differentials, medications and symptoms with fCal outcomes and infections. Correlation between HBV chronicity (among participants who attended both 2023 and 2024 follow-up timepoints) and HIV viral suppression (defined as viral load <1000 copies/mL) with fCal outcomes also was examined. Sensitivity analyses were conducted in which one participant with imputed malaria and HBV status was excluded from the models. They had missing RDT data for both infections, and their status was imputed as negative based on the absence of self-reported previous diagnosis in 2024, no positive HBV RDT in 2023, and negative malaria slide results in 2024.

**Biomedical variables**

WBC differentials (cell number x $10^9$/mL of blood) were collected from finger pricks (HemoCue WBC DIFF System). Stool samples were subject to a visual inspection by experienced technicians. Stool consistency was measured by the Bristol stool scale and stool types (from 1 to 7) were categorised as constipation (types 1 and 2), normal (types 3, 4 and 5), or diarrhoea (types 6 and 7) [6]. Technicians also recorded any visible blood in the stool and any indication of melena (black, tarry stool). Faecal occult blood (FOB), non-visible blood in stool, also was assessed (Vaxpert FOB Test). Nurses asked participants about gastrointestinal symptoms experienced in the past month, at first recording symptoms from a predefined list believed to be related to intestinal schistosomiasis (big belly, mass in abdomen, severe abdominal pain, vomiting blood, bloody diarrhoea, blood in stool without diarrhoea, black or charcoal stool, collapsed/fainting, jaundice/yellow eyes/yellow face), followed by an open answer question about any symptoms which were recorded as a free-text response. Biosample-based and self-reported symptoms were combined to create binary indicators: any gastrointestinal symptoms, any abdominal pain (general, stomach, lower, and severe, epigastric pain), abdominal enlargement (abdominal mass, distention, bloating, swelling, big tummy), vomiting or nausea, vomiting blood, FOB, any blood in stool (bloody diarrhoea or blood in stool without diarrhoea), melena, any diarrhoea without blood (self-reported diarrhoea or diarrhoea based on the Bristol stool scale), constipation (based on the Bristol stool scale). Nurses also asked about and any medication taken within the past month (coded as binary indicators), which included other deworming medications, antimalarials, antibiotics, and anti-inflammatory medications (corticosteroids or non-steroidal anti-inflammatory drugs). They also asked about praziquantel received through MDA within the past year and praziquantel received through the study on the day of the survey. We created a binary indicator of praziquantel received through the study within the past year based on the 2023 survey, and a combined indicator of praziquantel received within the past year (through MDA or the study). HIV counsellors recorded whether individuals self-reported initiating ART for HIV or HBV [7]. At the SchistoTrack 2025 annual follow-up timepoint, people living with HIV (PLHIV) had viral load measured, as described elsewhere [7].

**Table S1. Calprotectin cohort representativeness.** Comparison of age, gender and key exposure variables between the calprotectin cohort (N = 640) and the wider Pakwach district cohort (N = 1464).

| | Calprotectin cohort (N = 640) | Pakwach cohort (N = 1464) |
|---|---|---|
| **Age (mean)** | 25.6 | 25.8 |
| **Adults (≥18 years)** | 322 (50.3%) | 735 (50.2%) |
| **Children (<18 years)** | 318 (49.7%) | 729 (49.8%) |
| **Gender (female)** | 328 (51.3%) | 715 (48.8%) |
| **Male** | 312 (48.8%) | 749 (51.2%) |
| **Year of recruitment** | | |
| **2022** | 106 (16.6%) | 751 (51.3%) |
| **2023** | 470 (73.4%) | 537 (36.7%) |
| **2024** | 64 (10.0%) | 176 (12.0%) |
| ***S. mansoni* status (positive)** | 314 (49.1%) | 766 (52.3%) |
| **No infection** | 326 (50.9%) | 679 (46.4%) |
| **Low infection** | 132 (20.6%) | 322 (22.0%) |
| **Mild infection** | 107 (16.7%) | 241 (16.5%) |
| **High infection** | 75 (11.7%) | 203 (13.9%) |
| **Malaria status (positive)** | 236 (36.9%) | 565 (38.6%) |
| **HIV status (positive)** | 33 (5.2%) | 68 (4.7%) |
| **HIV status-aware** | 26 (78.8%) | 53 (77.9%) |
| **Ever HIV ART** | 26 (78.8%) | 53 (77.9%) |
| **Current HIV ART** | 26 (78.8%) | 52 (76.5%) |
| **HBV status-positive (2023)** | 29 (4.5%) | 89 (6.1%) |
| **HBV status-positive (2024)** | 34 (5.3%) | 85 (5.8%) |

**Table S2. Unadjusted odds ratios of detectable calprotectin (fCal).** Odds ratios from univariate logistic regression models of infection, sociodemographic, and water and sanitation covariates with detectable fCal.

| | OR (95% CI) | *p*-value |
|---|---|---|
| **S. mansoni status (positive)** | 2.13 (1.47–3.07) | <0.001 |
| **S. mansoni infection intensity (EPG)** | 1.003 (1.001–1.004) | <0.001 |
| **Malaria status (positive)** | 1.12 (0.77–1.63) | 0.546 |
| **Malaria parasite density** | 1.00 (1.00–1.00) | 0.581 |
| **HIV status (positive)** | 1.46 (0.59–3.62) | 0.415 |
| **HBV status (positive)** | 1.13 (0.50–2.54) | 0.774 |
| **Age** | 0.99 (0.98–1.00) | 0.193 |
| **Gender (female)** | 1.04 (0.73–1.48) | 0.849 |
| **Occupation** | | |
| **Farmer** | 1.18 (0.72–1.94) | 0.520 |
| **Fisherman** | 1.18 (0.49–2.80) | 0.716 |
| **Fishmonger** | 1.79 (0.21–15.45) | 0.597 |
| **Education** | | |
| **Primary** | 0.89 (0.56–1.41) | 0.630 |
| **Secondary or above** | 1.12 (0.49–2.52) | 0.790 |
| **Year of recruitment (ref. 2024)** | | |
| **2022** | 7.90 (2.77–22.59) | <0.001 |
| **2023** | 0.91 (0.51–1.63) | 0.758 |
| **No. of people in household** | 1.00 (0.88–1.14) | 0.975 |
| **Home quality** | 1.02 (0.95–1.11) | 0.580 |
| **Social status** | 0.98 (0.61–1.57) | 0.937 |
| **Home ownership** | 0.56 (0.27–1.16) | 0.120 |
| **Years lived in village** | 1.00 (0.99–1.01) | 0.492 |
| **Distance to nearest gov't health centre** | 1.00 (1.00–1.00) | 0.046 |
| **Primary type of health facility used (ref. Private or other)** | | |
| **Gov't health centre** | 0.45 (0.19–1.09) | 0.076 |
| **Drinking water (improved)** | 0.38 (0.26–0.56) | <0.001 |
| **Sanitation (improved)** | 2.00 (1.39–2.89) | <0.001 |
| **Treated water** | 1.37 (0.91–2.08) | 0.136 |

OR – odds ratio; CI – confidence interval.

**Table S3. Unadjusted odds ratios of calprotectin (fCal) ≥100 µg/g.** Odds ratios from univariate logistic regression models of infection, sociodemographic, and water and sanitation covariates with detectable fCal ≥100 µg/g.

| | OR (95% CI) | *p*-value |
|---|---|---|
| **S. mansoni status (positive)** | 1.43 (0.98–2.07) | 0.062 |
| **S. mansoni infection intensity (EPG)** | 1.00 (1.00–1.00) | <0.001 |
| **Malaria status (positive)** | 0.94 (0.64–1.39) | 0.767 |
| **Malaria parasite density** | 1.00 (1.00–1.00) | 0.692 |
| **HIV status (positive)** | 2.30 (1.09–4.87) | 0.029 |
| **HBV status (positive)** | 1.72 (0.82–3.61) | 0.154 |
| **Age** | 0.99 (0.98–1.00) | 0.026 |
| **Gender (female)** | 0.99 (0.68–1.44) | 0.956 |
| **Occupation** | | |
| **Farmer** | 0.65 (0.37–1.14) | 0.130 |
| **Fisherman** | 1.41 (0.63–3.16) | 0.408 |
| **Fishmonger** | 0.00 (0.00–Inf) | 0.981 |
| **Education** | | |
| **Primary** | 0.86 (0.54–1.36) | 0.523 |
| **Secondary or above** | 0.55 (0.22–1.36) | 0.198 |
| **Year of recruitment (ref. 2024)** | | |
| **2022** | 0.94 (0.47–1.91) | 0.875 |
| **2023** | 0.74 (0.41–1.34) | 0.318 |
| **No. of people in household** | 1.00 (0.87–1.15) | 0.993 |
| **Home quality** | 0.97 (0.89–1.05) | 0.418 |
| **Social status** | 1.27 (0.79–2.05) | 0.328 |
| **Home ownership** | 0.90 (0.47–1.73) | 0.750 |
| **Years lived in village** | 1.00 (0.99–1.01) | 0.944 |
| **Distance to nearest gov't health centre** | 1.00 (1.00–1.00) | 0.035 |
| **Primary type of health facility used (ref. Private or other)** | | |
| **Gov't health centre** | 0.83 (0.41–1.68) | 0.598 |
| **Drinking water (improved)** | 0.75 (0.52–1.09) | 0.131 |
| **Sanitation (improved)** | 1.31 (0.90–1.90) | 0.160 |
| **Treated water** | 0.84 (0.55–1.29) | 0.435 |

OR – odds ratio; CI – confidence interval.

**Table S4. Unadjusted odds ratios of calprotectin (fCal) >250 µg/g.** Odds ratios from univariate logistic regression models of infection, sociodemographic, and water and sanitation covariates with fCal >250 µg/g.

| | OR (95% CI) | *p*-value |
|---|---|---|
| **S. mansoni status (positive)** | 1.97 (1.05–3.71) | 0.035 |
| **S. mansoni infection intensity (EPG)** | 1.00 (1.00–1.00) | 0.035 |
| **Malaria status (positive)** | 0.76 (0.40–1.47) | 0.419 |
| **Malaria parasite density** | 1.00 (1.00–1.00) | 0.958 |
| **HIV status (positive)** | 2.05 (0.69–6.15) | 0.199 |
| **HBV status (positive)** | 1.84 (0.62–5.47) | 0.274 |
| **Age** | 0.98 (0.96–1.00) | 0.028 |
| **Gender (female)** | 2.22 (1.16–4.26) | 0.016 |
| **Occupation** | | |
| **Farmer** | 0.80 (0.33–1.95) | 0.622 |
| **Fisherman** | 1.44 (0.42–4.96) | 0.568 |
| **Fishmonger** | 0.00 (0.00–Inf) | 0.989 |
| **Education** | | |
| **Primary** | 1.16 (0.53–2.58) | 0.708 |
| **Secondary or above** | 0.67 (0.14–3.30) | 0.627 |
| **Year of recruitment (ref. 2024)** | | |
| **2022** | 0.81 (0.31–2.14) | 0.671 |
| **2023** | 0.41 (0.18–0.95) | 0.037 |
| **No. of people in household** | 0.88 (0.69–1.13) | 0.309 |
| **Home quality** | 1.06 (0.94–1.18) | 0.337 |
| **Social status** | 0.90 (0.39–2.07) | 0.806 |
| **Home ownership** | 0.47 (0.20–1.10) | 0.081 |
| **Years lived in village** | 1.00 (0.98–1.02) | 0.815 |
| **Distance to nearest gov't health centre** | 1.00 (1.00–1.00) | 0.259 |
| **Primary type of health facility used (ref. Private or other)** | | |
| **Gov't health centre** | 3.34 (0.45–24.86) | 0.239 |
| **Drinking water (improved)** | 0.44 (0.23–0.84) | 0.012 |
| **Sanitation (improved)** | 1.87 (1.00–3.48) | 0.050 |
| **Treated water** | 1.24 (0.64–2.39) | 0.523 |

OR – odds ratio; CI – confidence interval.

**Table S5. Unadjusted odds ratios of calprotectin (fCal) concentration.** Odds ratios from univariate linear regression models of infection, sociodemographic, and water and sanitation covariates with fCal concentration.

| | OR (95% CI) | *p*-value |
|---|---|---|
| **S. mansoni status (positive)** | 0.19 (-0.03–0.42) | 0.096 |
| **S. mansoni infection intensity (EPG)** | 0.001 (0.001–0.001) | <0.001 |
| **Malaria status (positive)** | 0.06 (-0.18–0.29) | 0.633 |
| **Malaria parasite density** | 0.00 (-0.00–0.00) | 0.627 |
| **HIV status (positive)** | 0.39 (-0.11–0.89) | 0.130 |
| **HBV status (positive)** | 0.30 (-0.20–0.79) | 0.239 |
| **Age** | -0.01 (-0.01–-0.00) | 0.022 |
| **Gender (female)** | -0.06 (-0.28–0.16) | 0.603 |
| **Occupation** | | |
| **Farmer** | -0.23 (-0.53–0.08) | 0.142 |
| **Fisherman** | 0.22 (-0.31–0.74) | 0.416 |
| **Fishmonger** | -1.01 (-2.10–0.09) | 0.073 |
| **Education** | | |
| **Primary** | -0.02 (-0.31–0.26) | 0.876 |
| **Secondary or above** | -0.06 (-0.55–0.42) | 0.795 |
| **Year of recruitment (ref. 2024)** | | |
| **2022** | -0.16 (-0.60–0.27) | 0.467 |
| **2023** | -0.31 (-0.69–0.08) | 0.120 |
| **No. of people in household** | -0.05 (-0.13–0.04) | 0.262 |
| **Home quality** | -0.02 (-0.06–0.03) | 0.501 |
| **Social status** | 0.09 (-0.21–0.39) | 0.553 |
| **Home ownership** | 0.07 (-0.31–0.46) | 0.713 |
| **Years lived in village** | -0.00 (-0.01–0.00) | 0.682 |
| **Distance to nearest gov't health centre** | -0.00 (-0.00–-0.00) | 0.036 |
| **Primary type of health facility used (ref. Private or other)** | | |
| **Gov't health centre** | 0.06 (-0.36–0.48) | 0.776 |
| **Drinking water (improved)** | -0.08 (-0.30–0.15) | 0.511 |
| **Sanitation (improved)** | 0.13 (-0.09–0.36) | 0.240 |
| **Treated water** | -0.13 (-0.38–0.12) | 0.313 |

OR – odds ratio; CI – confidence interval.

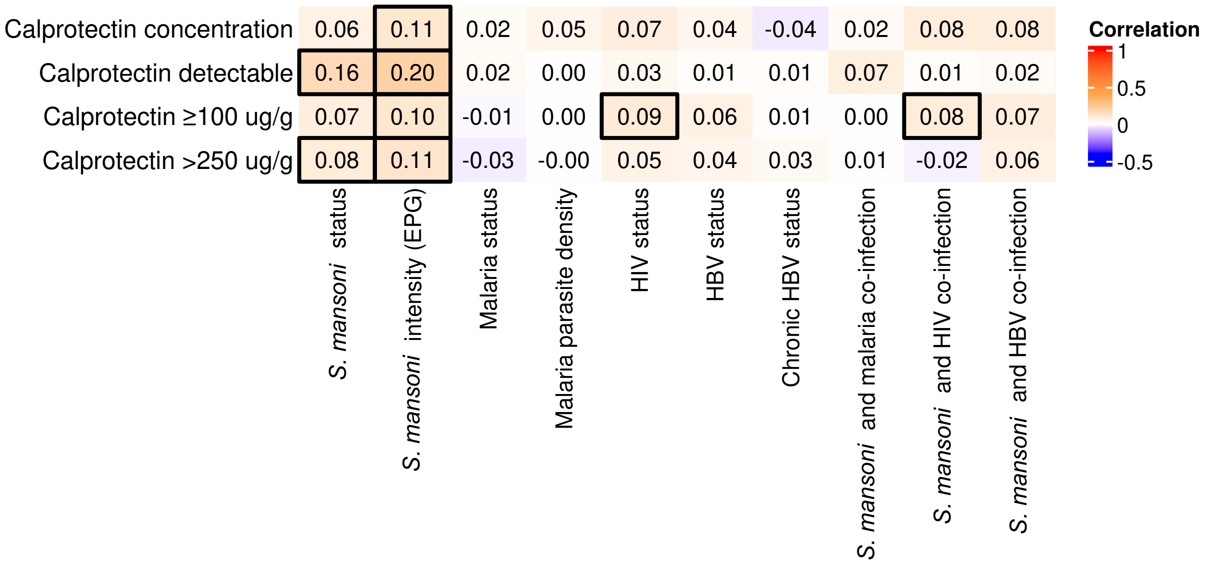

**Figure S1.** Spearman correlations of single and co-infections with calprotectin (fCal).

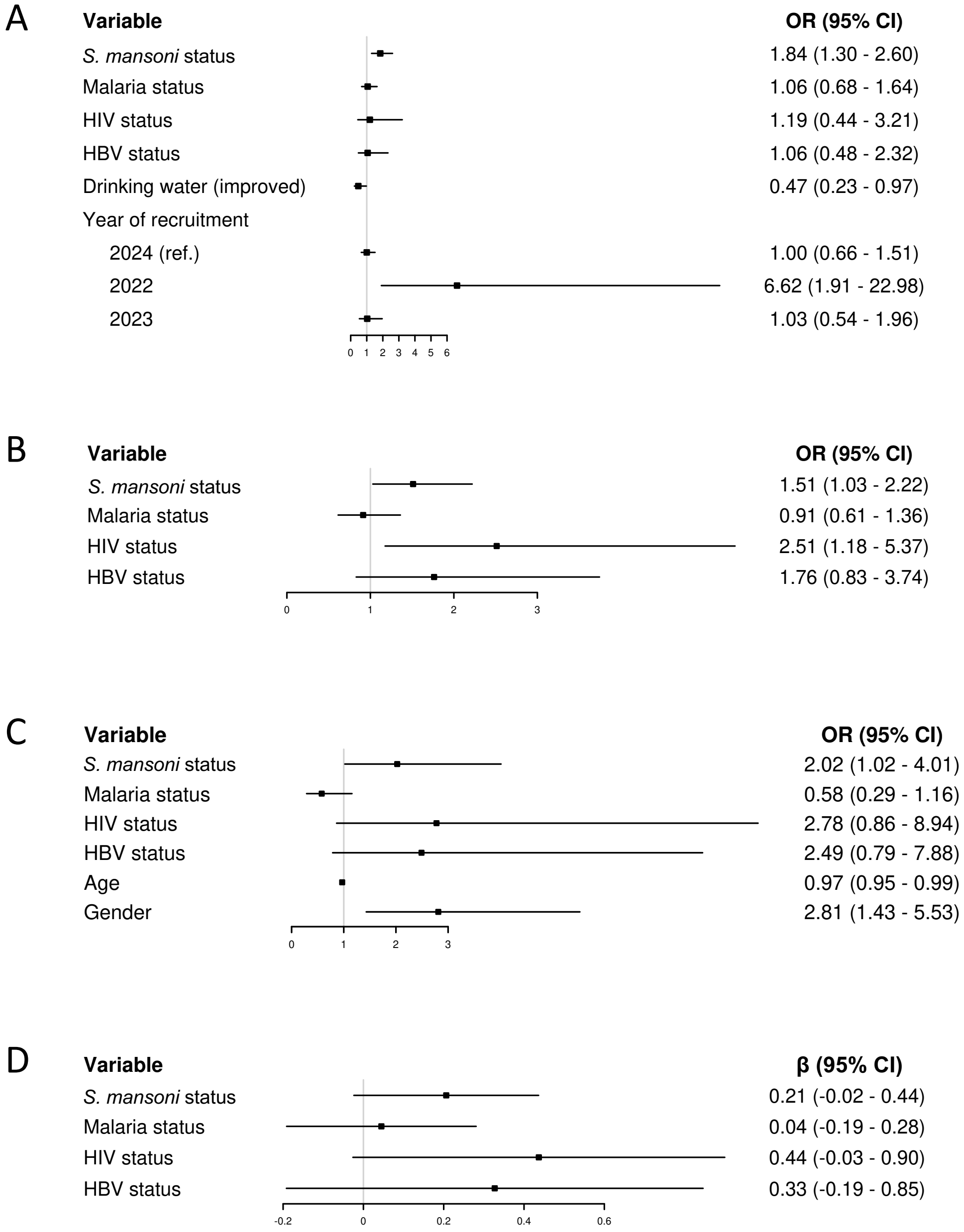

**Figure S2. Calprotectin (fCal) models for infection status.** *S. mansoni* infection intensity and malaria parasite density were replaced with *S. mansoni* and malaria status. Model of (A) detectable fCal, (B) ≥100 µg/g, (C) fCal >250 µg/g, and (D) linear fCal (N = 640). HIV status remained significant for fCal ≥100 µg/g. Malaria and HBV remained non-significant in all models. The year of recruitment became significant; participants recruited in 2022 had 6.62 (95% CI 1.91-22.98) times higher odds of having detectable fCal compared to participants recruited in 2024. Improved drinking water source remained significant, while improved sanitation was no longer selected. Age was additionally selected in the fCal >250 µg/g model, with each additional year lowering the odds of high levels of gut inflammation by 0.03 (OR 0.97, 95% CI 0.94-0.99). Gender remained significant. Logistic regression models were selected by backward stepwise selection based on the lowest Bayesian Information Criterion. 95% confidence intervals were calculated using clustered standard errors at the village level (for detectable fCal; number of villages = 12) and the household level (for linear fCal; number of households = 376). Floating absolute risks were calculated for the year of recruitment variable. ICC (village) = 0.147; 0.029; 0.014; 0.016. ICC (household) = 0.119; 0.062; 0.084; 0.102. VIF range: 1.00-1.05; 1.01-1.04; 1.05-1.27; 1.01-1.03. AUC for stratified ten-fold cross-validation = 0.66; 0.55; 0.70. $R^2$ (linear model) = 0.02.

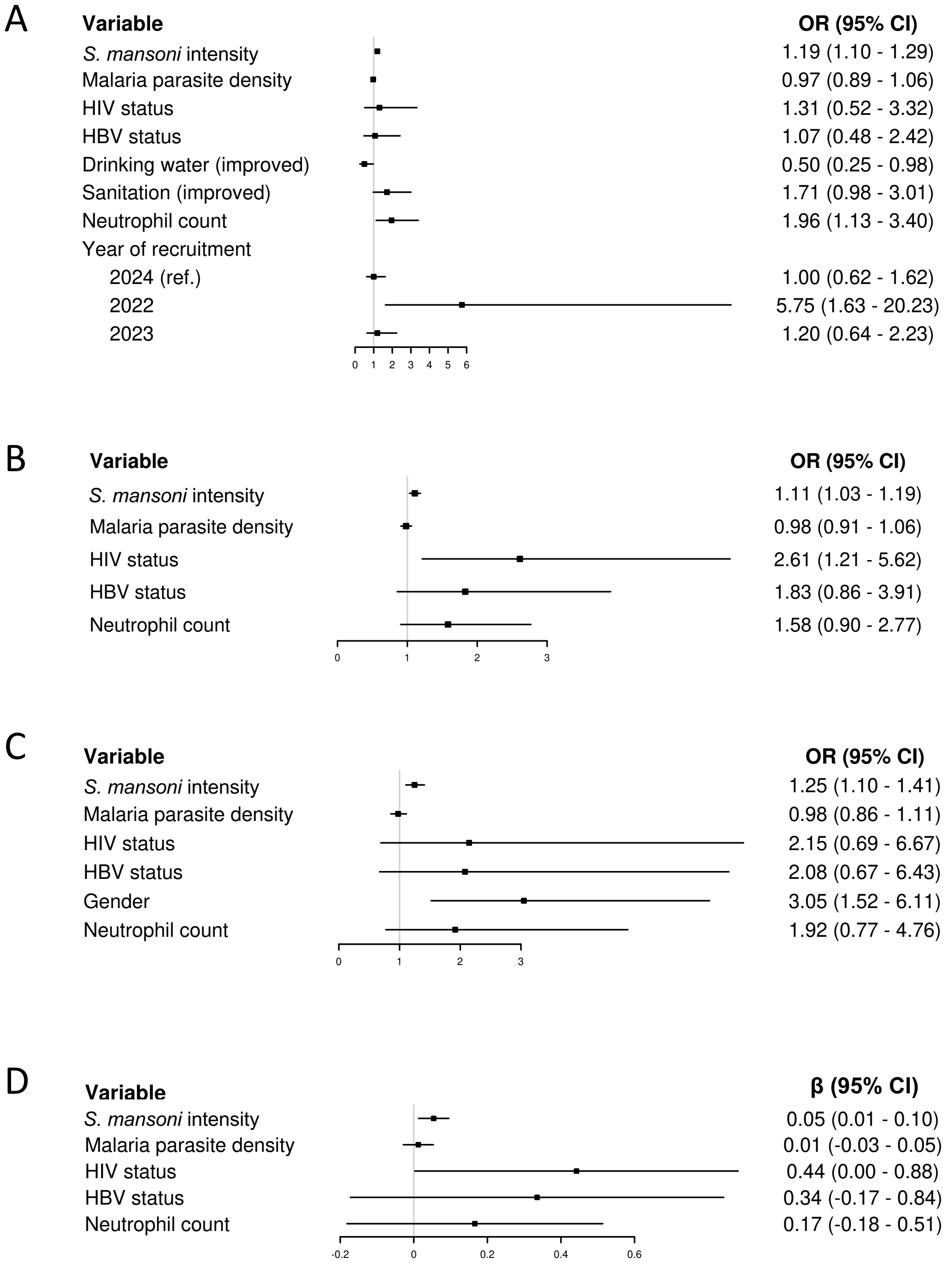

**Figure S3. Mediator analysis of neutrophils (infection intensity).** Calprotectin (fCal) models for infection intensity including natural log-transformed neutrophil count as a potential mediator. Model of (A) detectable fCal, (B) ≥100 µg/g, (C) fCal >250 µg/g, and (D) linear fCal (N = 640). Logistic regression models were selected by backward stepwise selection based on the lowest Bayesian Information Criterion. 95% confidence intervals were calculated using clustered standard errors at the village level (for detectable fCal; number of villages = 12) and the household level (for linear fCal; number of households = 376). Floating absolute risks were calculated for the year of recruitment variable.

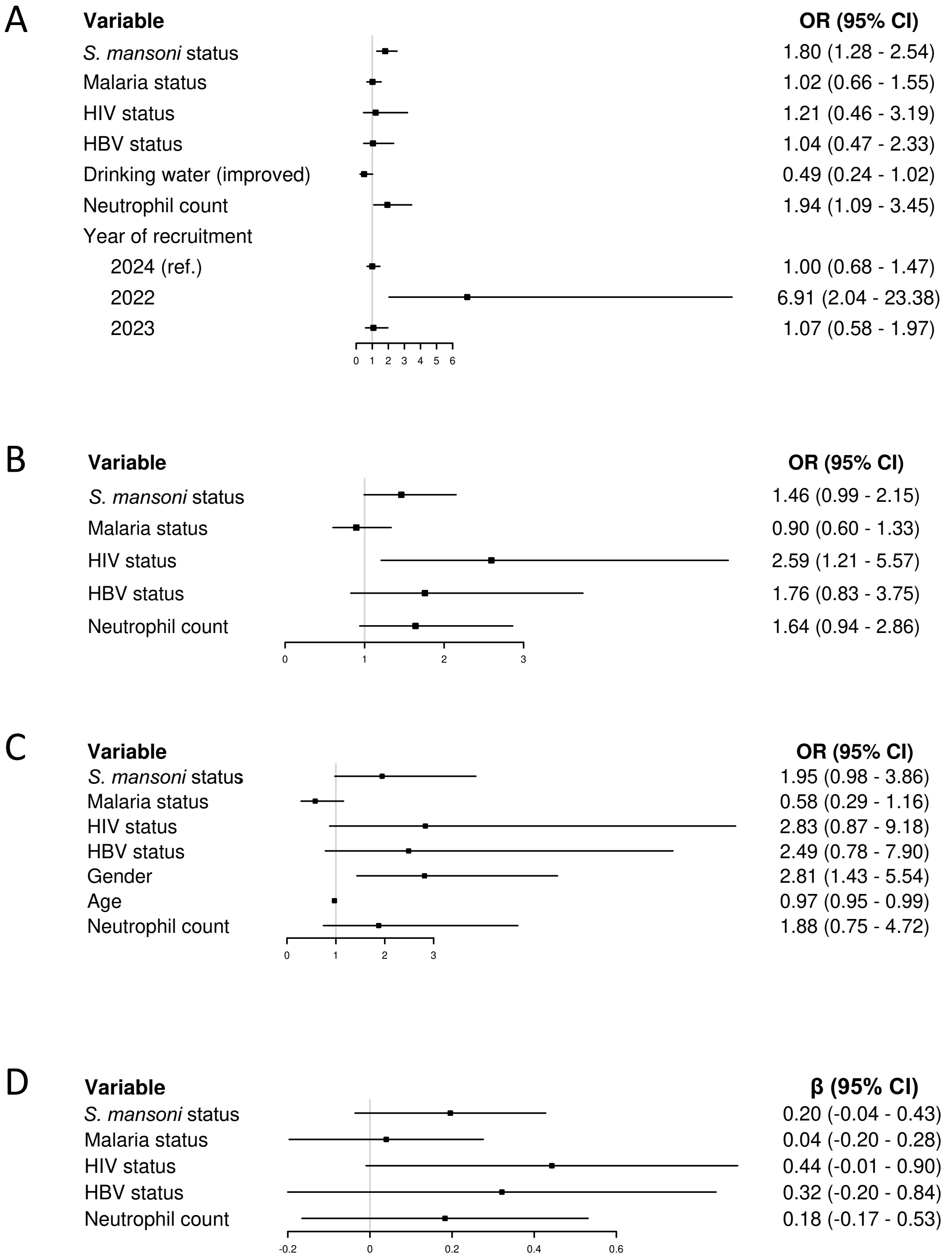

**Figure S4. Mediator analysis of neutrophils (infection status).** Calprotectin (fCal) models for infection status including natural log-transformed neutrophil count as a potential mediator. Model of (A) detectable fCal, (B) ≥100 μg/g, (C) fCal >250 μg/g, and (D) linear fCal (N = 640). Logistic regression models were selected by backward stepwise selection based on the lowest Bayesian Information Criterion. 95% confidence intervals were calculated using clustered standard errors at the village level (for detectable fCal; number of villages = 12) and the household level (for linear fCal; number of households = 376). Floating absolute risks were calculated for the year of recruitment variable.

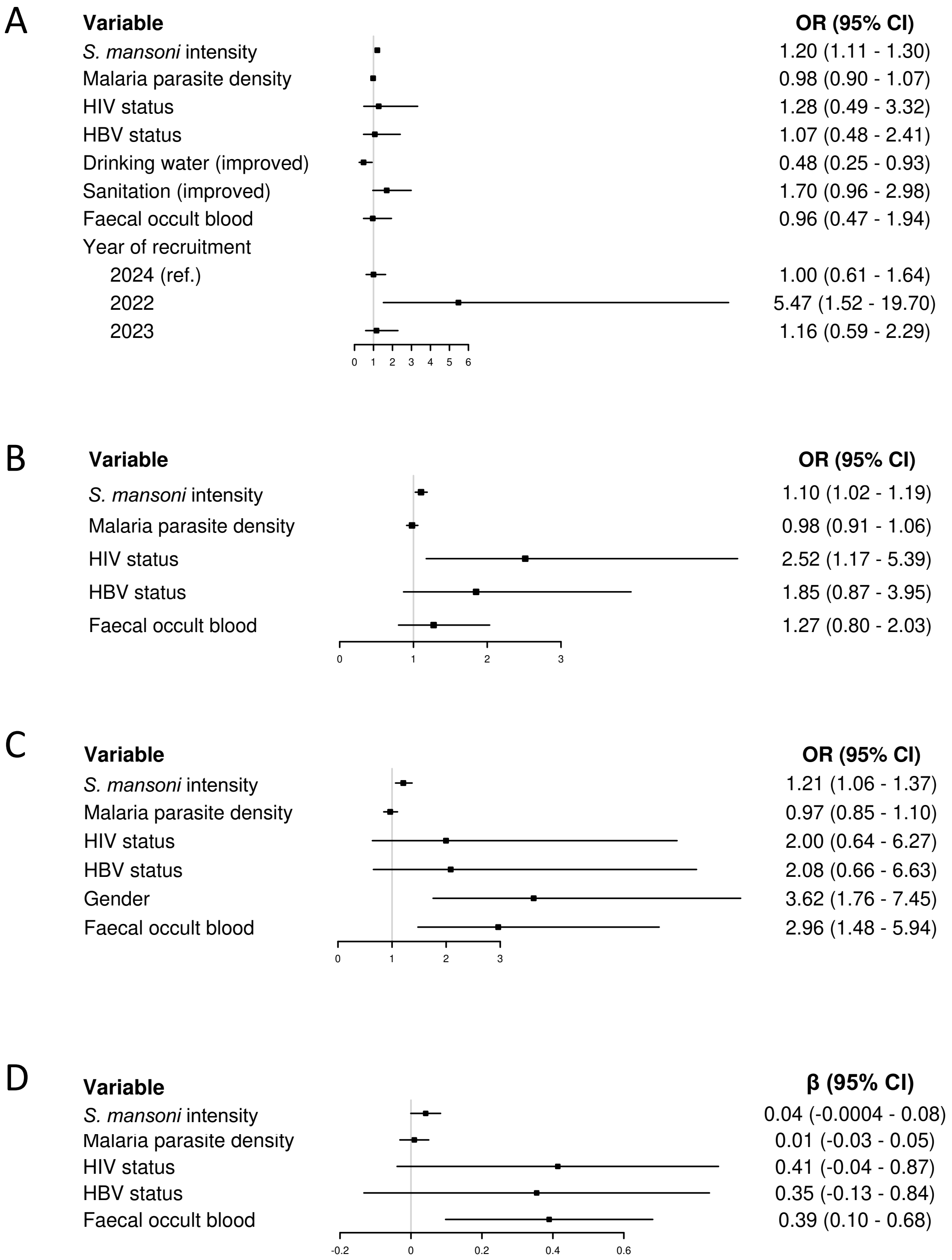

**Figure S5. Mediator analysis of faecal occult blood (infection intensity).** Calprotectin (fCal) models for infection intensity including faecal occult blood as a potential mediator. Model of (A) detectable fCal, (B) ≥100 µg/g, (C) fCal >250 µg/g, and (D) linear fCal (N = 640). Logistic regression models were selected by backward stepwise selection based on the lowest Bayesian Information Criterion. 95% confidence intervals were calculated using clustered standard errors at the village level (for detectable fCal; number of villages = 12) and the household level (for linear fCal; number of households = 376). Floating absolute risks were calculated for the year of recruitment variable.

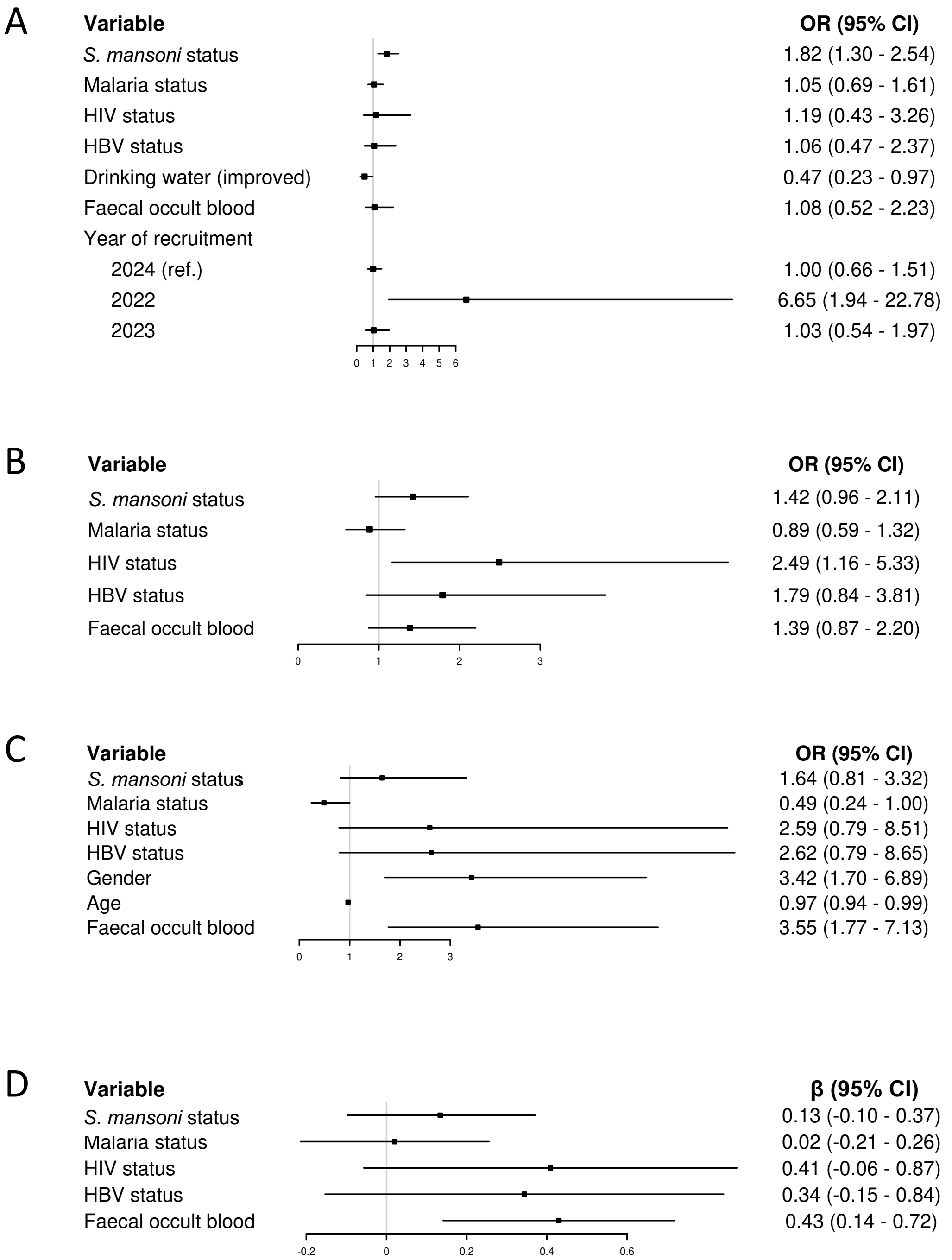

**Figure S6. Mediator analysis of faecal occult blood (infection status).** Calprotectin (fCal) models for infection status including faecal occult blood as a potential mediator. Model of (A) detectable fCal, (B) ≥100 µg/g, (C) fCal >250 µg/g, and (D) linear fCal (N = 640). Logistic regression models were selected by backward stepwise selection based on the lowest Bayesian Information Criterion. 95% confidence intervals were calculated using clustered standard errors at the village level (for detectable fCal; number of villages = 12) and the household level (for linear fCal; number of households = 376). Floating absolute risks were calculated for the year of recruitment variable.
